# Facilitating the Measurement and Treatment of Behavioral and Psychological Symptoms of Dementia (BPSD) and Understanding Caregiver Burden Using Wearable Devices in Rural Taiwan – Protocol for a Dyadic Feasibility Pilot Study

**DOI:** 10.64898/2026.01.20.26344458

**Authors:** Ta-Wei Guu, Wan-Jing Li, Shih-Hsiung Lee, Chih-Shan Hsu, Chia-Ni Chou, Leon Lack, Wei-Fen Ma

## Abstract

**Introduction:** Alzheimer’s disease (AD) prevalence rises with societal ageing. In clinical care, behavioral and psychological symptoms of dementia (BPSD)—including depression, agitation/aggression, apathy, and sleep disturbance—worsen patients’ quality of life and substantially increase caregiver burden, more significantly than the cognitive symptoms. Standard BPSD assessments rely on caregiver-rated questionnaires that are cross-sectional and may be biased when caregivers are themselves older adults. Device-based measures (e.g., research-grade wrist actigraphy) can provide objective longitudinal data and novel features. In parallel, therapeutic wearables may improve sleep and mood in adults, and might improve BPSD if accepted by people living with dementia. This study aims to assess the feasibility and acceptability of two wearables (Geneactiv actigraphy and Re-Timer circadian regulator) among AD patients with significant BPSD and their caregivers in Taiwan.

**Methods:** This dyadic pilot study will recruit 20 participants (n=10 AD patients; n=10 caregivers) from outpatient services and affiliated day-care/dementia hubs in rural Taiwan. Participants will wear Geneactiv continuously for 8 weeks and Re-Timer ≥30 min/day for 4 weeks. Device-based data will be processed with GGIR, a well validated R-package designed for processing accelerometer data. Questionnaire assessments include Pittsburgh Sleep Quality Index, PSQI (PSQI), Neuropsychiatry Inventory Questionnaire (NPI-Q), Caregiver Burden Inventory (CBI), and a semi-structured interview based on the Taiwanese version of Quebec User Evaluation of Satisfaction with Assistive Technology (T-QUEST) at prespecified timepoints.

**Discussion:** Wearable devices may facilitate the measurement and treatment of specific BPSD, as well as reduce caregiver burden. If proven feasible even in rural Taiwan where both digital and health literacy and resources are limited, this model will inform how device-based dementia care model can be considered and applied in the context of global ageing.

**Ethics & registration:** This protocol has been approved by the China Medical University and Hospital Research Ethics Committee (CMUH114-REC3-072), and pre-registered in ClinicalTrials.gov (NCT07249918).

## 1. Introduction

Globally, the number of people living with dementia has been projected to increase to 153 million by 2050 [1], and will result in huge societal cost. As Taiwan entering a “super-aged society” (≥20 percent of the population aged ≥65 years old), the rapidly rising AD cases and reduced younger population substantially increase care demands, especially in rural areas [2]. AD patients frequently present with behavioural and psychological symptoms (BPSD) [3, 4]. Among all the BPSD symptoms, depression, agitation/aggression, apathy, and nocturnal sleep problems often consume the most care resources and markedly elevate caregiver stress and sleep disturbances [5, 6].

Current assessment of BPSD depends on clinical interviews and caregiver-completed questionnaires such as the Neuropsychiatry Inventory (NPI) and Neuropsychiatry Inventory Questionnaire (NPI-Q) [7, 8], which may be imprecise given caregiver age-related declines in memory and verbal function, especially in a super-aged society context. Device-based measurement (using research-grade actigraphy such as Geneactiv) can capture objective, longitudinal activity, sleep, and light-exposure patterns [9–11], and its derived metrics (e.g., sleep regularity index, SRI) have clinical associations with both physical and mental health outcomes [12–14].

On the treatment side, pharmacotherapy remains a common modality for BPSD despite limited approvals and known risks such as falls and mortality [15]. Non-pharmacological approaches are therefore important and are recommended by guidelines as the first line treatment [16, 17]. Light plays an important role in human life, and can be delivered using wearable devices with therapeutic potential [18]. Bright-light interventions, for example, when delivered via ambient systems show mixed efficacy in people living with dementia, potentially due to distance from the eye and reduced light reception capacity in this population [19]. Re-Timer is a wearable light-emitting circadian regulating goggle (500-nm blue-green light) with preliminary evidence for effects on mood, circadian phase, and sleep in adults and older adults [20–22]. If acceptable to people living with dementia and their caregivers, it may alleviate BPSD and reduce caregiver burden.

To the best of our knowledge, no previous studies have investigated the feasibility of using wearable devices for both measuring and treating BPSD, and using the same devices to monitor and alleviate the caregiver burden in parallel. Our study aims to apply a mixed-methods approach to assess the feasibility, acceptability, and preliminary efficacy of Geneactiv (a well-validated research-grade actigraphy[9, 23, 24]) and Re-Timer for AD patients and their caregiver. Here we describe the design, participants, intervention and outcome measurements of the study.

## 2. Materials and methods

### 2.1 Objectives

- Primary: Determine the acceptability and feasibility of Geneactiv and Re-Timer in AD patients with significant BPSD and their caregivers.
- Secondary: Obtain preliminary information on wearables’ influence on BPSD and sleep; examine relationships between light exposure, circadian metrics, and BPSD; compare device-based vs questionnaire-based measurements to support future digital biomarkers characterisation.

### 2.2 Study design and setting

This is a single-arm, dyadic pilot feasibility study lasting12 weeks, and has started recruitment on 17^th^ November 2025 in CMU Beigang Hospital outpatient psychiatry and neurology clinics, and affiliated dementia day-care center and hubs. Dyads comprise an AD/ mild cognitive impairment (MCI) due-to-AD patient with significant BPSD, and their adult primary caregiver. The recruitment and data collection will be completed in May and August 2026, respectively, while the results are expected to be available in December 2026 (see Figure 1 for the overall timeline).

**Figure 1.**
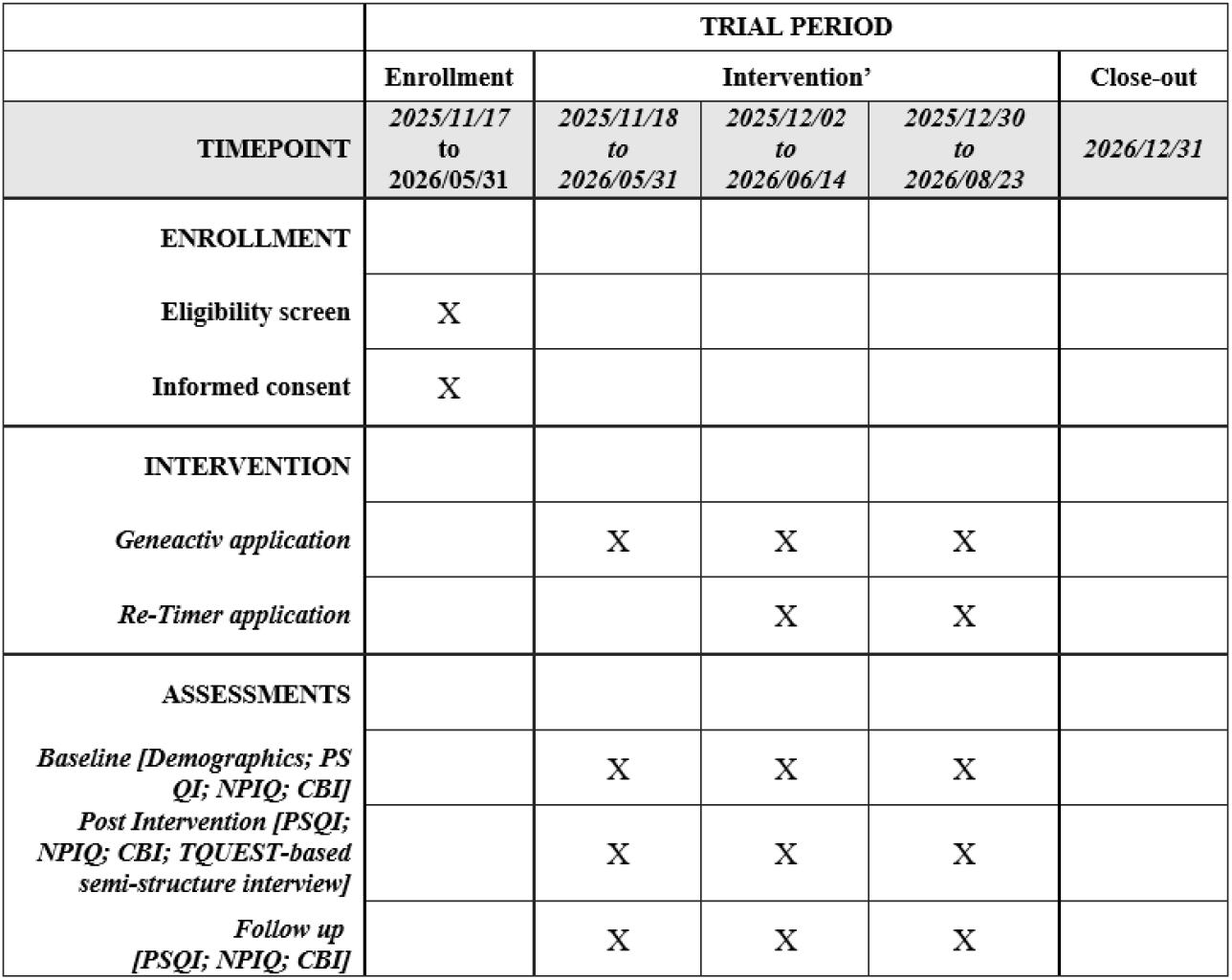
Timeline of the study. Abbreviations: PSQI=Pittsburgh Sleep Quality Index; NPIQ=Neuropsychiatric Inventory Questionnaire; CBI=Caregiver Burden Inventory; TQUEST=Taiwanese Version of the Quebec User Evaluation of Satisfaction with Assistive Technology.

#### Eligibility criteria

##### Inclusion criteria for patients (probable AD or MCI due to AD)

- Clinical diagnosis of probable AD or MCI due to AD (CDR 0.5) per the National Institute on Aging and Alzheimer’s Association (NIA-AA) diagnostic guidelines [25].
- At least one significant BPSD (depression, agitation/aggression, apathy, nocturnal sleep disturbance) defined as any Neuropsychiatry Inventory item score ≥4 (frequency x severity) [8] or Neuropsychiatry Inventory Questionnaire (NPI-Q) item severity ≥2 [26]. If sleep is the main symptom, Pittsburgh Sleep Quality Index, PSQI (PSQI) global score >5 [27].
- Stable clinical setting/environment ≥2 weeks (community residence/day-care engagement).
- Stable BPSD treatment (pharmacologic/non-pharmacologic) ≥2 weeks prior to enrollment.
- Able to provide a written informed consent or a legal representative consent.

##### Caregivers

- Adult primary caregiver of the enrolled patient.
- Stable treatments for caregiver stress (if any) ≥2 weeks prior to enrollment.
- Able to provide a written informed consent.

##### Exclusion criteria (patient/caregiver)

- Conditions increasing risk from bright-light exposure, including:
  - retinal disease;
  - current photosensitizing medications;
  - recent ocular surgery (<4 weeks) not fully recovered;
  - other conditions contraindicating blue-green light (e.g., epilepsy).
- Clinically unstable (e.g., in acute delirium; acute respiratory infection including COVID-19).

### 2.3. Assessments

#### Basic information collection

- AD Patients: Date of birth, sex, marital status, education level, time of dementia (or MCI) diagnosis, current cognitive status (based on the most recent assessment within six months using Clinical Dementia Rating (CDR), Cognitive Abilities Screening Instrument (CASI), or Mini-Mental State Examination (MMSE) (if recent cognitive changes are suspected, reassessment will be performed), time of onset of BPSD, current dementia-related medications, smoking status, alcohol use, medical and psychiatric history, and current treatments.
- Caregivers: Date of birth, sex, marital status, education level, smoking status, alcohol use, medical and psychiatric history, and current treatments. If the caregiver is an older adult with possible cognitive decline, their cognitive status will also be collected (CDR, CASI, or MMSE within six months; reassessment if recent changes are suspected).

#### Questionnaire Assessments

- **Pittsburgh Sleep Quality Index (PSQI) [27]:** A questionnaire assesses seven domains related to sleep. Each item scored 0 (no symptom) to 3. PSQI will be administered five times (Day 1, Day 15, Day 42, Day 56, Day 84) to both patients and caregivers. If the patient can complete the questionnaire independently, they will be advised to do so; otherwise, the caregiver will respond on their behalf.
- **Neuropsychiatric Inventory Questionnaire (NPI-Q) [26]:** Evaluates 12 BPSD symptoms. For each symptom: presence, severity (impact on patient, scored 1–3), and caregiver distress (impact on caregiver, scored 0–5) will be evaluated. It will be administered five times (Day 1, Day 15, Day 42, Day 56, Day 84) by the same primary caregiver throughout the study.
- **Caregiver Burden Inventory (CBI) [28]:** Assesses caregiver burden with 24 items across five domains (each scored 0–4); higher scores indicate greater burden. It will be administered three times (Day 1, Day 42, Day 84) by the same primary caregiver.
- **Taiwanese Version of the Quebec User Evaluation of Satisfaction with Assistive Technology (T-QUEST) [29]:** The T-QUEST will assesses satisfaction with wearables and related services. There are 13 items scored from 1 (very dissatisfied) to 5 (very satisfied), and will be administered to both sides of the dyads at Day 42 (for Re-Timer) and Day 56 (for Geneactiv). If the patient cannot complete the questionnaire, only the caregiver will respond.

#### Semi-Structured Interviews

The interviews will extend from T-QUEST topics to explore participants’ and caregivers’ experiences with both devices, including acceptability, ease of use, comfort, and challenges. Open questions will first be used, followed by iterative, in-depth probing that focuses on convenience, regularity, motivation, and satisfaction. Through this back-and-forth process, participants will be encouraged to provide everyday-life examples so that they can fully articulate their experiences and feelings when using the devices. Each interview will last approximately 30–60 minutes in a setting comfortable for participants and caregivers to ensure natural and reliable data collection. Audio recordings will be transcribed and analyzed using thematic analysis to explore perceptions of wearable devices for assessing and managing neuropsychiatric symptoms after trial participation. See supplementary materials for the interview guide.

### 2.4. Device and interventions procedures

#### Geneactiv wrist-worn actigraphy

Geneactiv is a watch-like device capturing physical activities with a three-axis accelerometer (Figure 2, left). A light sensor on its surface is capable of detecting ambient illuminance. It’s feasibility and acceptability have been previously validated in dementia populations by ours and other research group independently. Based on our previous results, the team will fit devices, offer manufacturer-approved alternative straps to optimize comfort/appearance, and instruct participants to avoid covering the light sensor.

**Figure 2.**
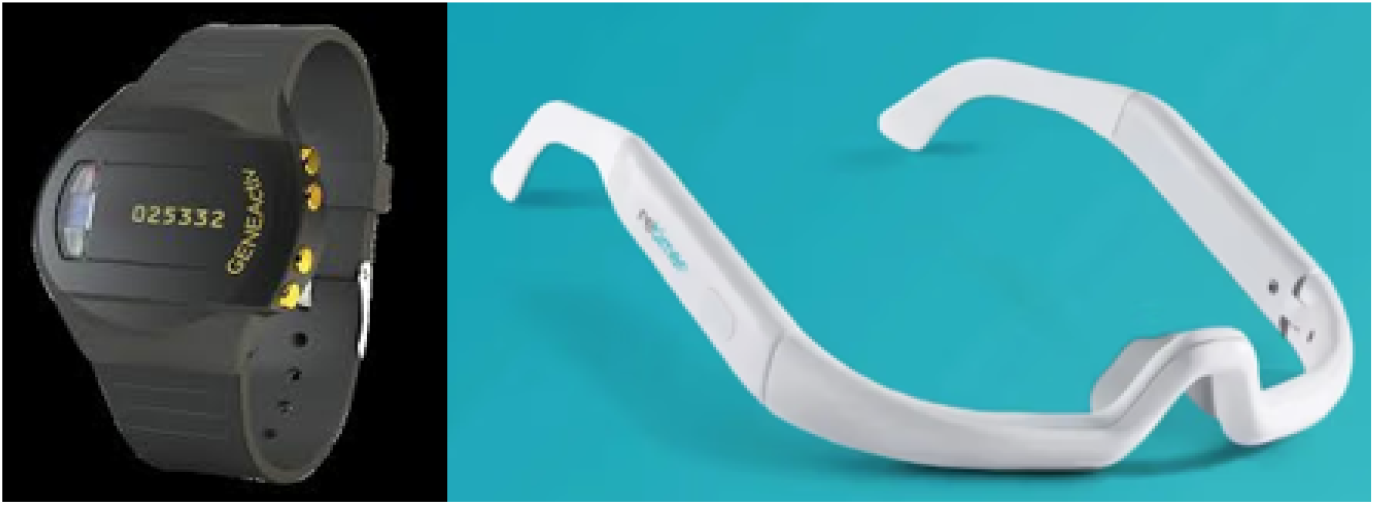
The two study devices. Left: Geneactiv research-grade actigraphy; right: Re-Timer circadian regulator.

#### Re-Timer circadian regulator

Re-Timer is an eyewear emitting 500-nm blue-green visible light with no ultraviolet hazard (Figure 2, right). The device power/irradiance: 506 lux, 230 μW/cm^2^. Theoretically, the light emitted could reduce melatonin secretion and enhance alertness. Prior studies support applicability and safety in adults, those with certain psychiatric conditions, and older adults.

#### Device-wearing and assessment schedule

Participants will be retained in the study for 12 weeks (as in Figure 3), during which they will be advised to continue their usual daily routine/care, and will be advised to wear the device for up to 8 weeks, based on the following schedule:

**Figure 3.**
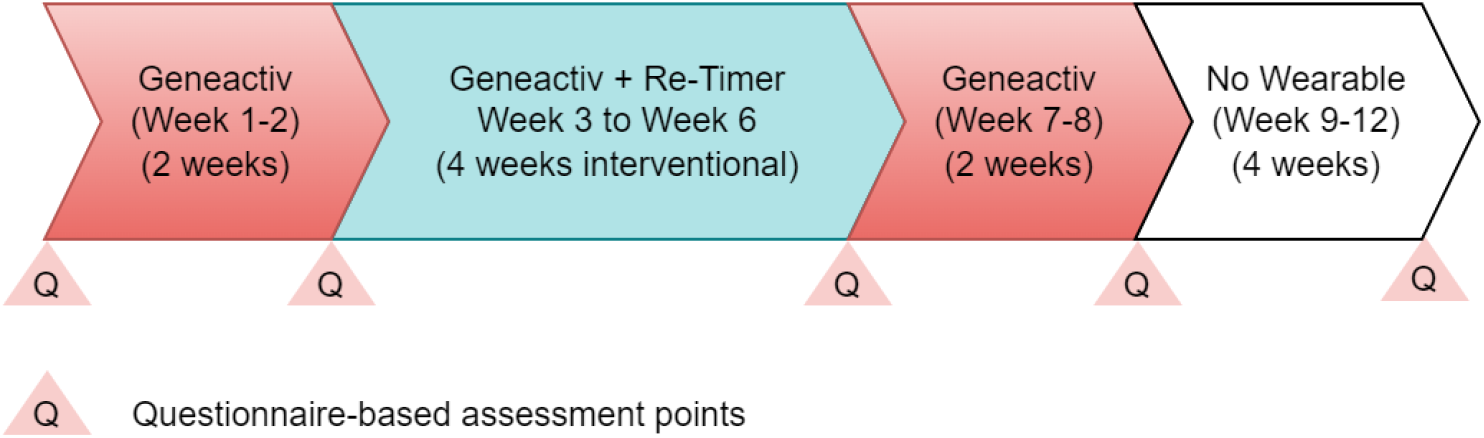
Study flow diagram. The study lasts for 12 weeks. Participants will be advised to wear the Geneactiv wrist actigraphy for up to 8 weeks, and the Re-Timer circadian regulator for up to 4 weeks. They will complete five rounds of assessments on Day 1 (baseline before any device-wearing), Day 15 (baseline before Re-Timer), Day 42 (immediate post-Re-Timer), Day 56 (post-Re-Timer and Geneactiv), and Day 84 (trial completion).

- **Day 1**: Baseline demographics; PSQI; NPI-Q; CBI; **Geneactiv start**.
- **Day 15**: PSQI; NPI-Q; **Re-Timer start** (daily 30–60 min).
- **Day 42**: PSQI; NPI-Q; CBI; T-QUEST-based semi-structure interview for Re-Timer; **Re-Timer end**.
- **Day 56**: PSQI; NPI-Q; T-QUEST-based semi-structure interview for Geneactiv; **Geneactiv end**.
- **Day 84**: Follow-up PSQI; NPI-Q; CBI.

More specifically on the device-wearing:

- **Geneactiv**: Days 1–56 (8 weeks) continuous wear. The device is waterproof, but participants may remove the device anytime during the trial if discomfort occurs.
- **Re-Timer**: Days 15–42 (4 weeks) ≥30 min each time, and up to 60 min/day. The participants will be recommended to wear it twice a day, one in the morning, one in early afternoon, but can remove the device and stop the treatment anytime if discomfort occurs. Day-care/hub participants may choose on-site staff-assisted wear and/or home wear. Correct positioning will be taught by the study team members. A participant manual will be provided to record the device-wearing time windows each day and any feedback.

## 3. Outcomes assessments and data analyses

### 3.1 Primary endpoints (feasibility/acceptability)

- **Wear compliance**: Geneactiv actual wear time (GGIR-derived); Re-Timer daily wear timing and duration.
- **Satisfaction (T-QUEST)** at day 42 (Re-Timer) and day 56 (Geneactiv). Between-group differences (patients vs caregivers) will be calculated by independent-samples t-tests.
- **Qualitative acceptability**: Semi-structured interviews probing convenience, comfort, barriers, and perceived value extended from the T-QUEST will be used. Transcription and thematic analysis with triangulation and independent coder checks will be done.

### 3.2 Secondary endpoints (exploratory)

- **BPSD (NPI-Q)** and **sleep (PSQI)** for patients and caregivers across five timepoints (days 1, 15, 42, 56, 84). **Caregiver Burden Inventory (CBI)** at days 1, 42, and 84.
- Device-based features: activity metrics; sleep metrics (sleep onset, awakenings, wake time, total sleep time, **SRI**); **light exposure** metrics; paired with NPI-Q/PSQI in time-series analyses to explore relationships among **light, circadian rhythm, and BPSD**.

### 3.3 Sample size and data analysis

This study is not powered for efficacy and will focus on descriptive feasibility outcomes. Therefore, a convenience sample targeted at 10 patient–caregiver dyads is planned.

Analyses will be conducted in R/RStudio for quantitative data:

- **Primary (feasibility/acceptability)**: Descriptive summaries of wear-time compliance and T-QUEST totals/subscales; between-group comparisons (patients versus caregivers) via independent-samples t-tests as appropriate.
- **Secondary (exploratory)**: GGIR extraction of activity, sleep (including sleep regularity index, SRI), and light; mixed-effects models for longitudinal repeated measures with covariates (age, disease severity, comorbidities). Evaluate potential dose–response between light intensity/duration and BPSD/sleep outcomes. Compute preliminary effect sizes (Cohen’s d) to inform future sample size planning. No formal efficacy hypothesis testing.
- **Qualitative data analysis**: They will begin concurrently with data collection. Throughout the analytic process, peer debriefing with experienced qualitative researchers will be conducted to critically review emerging codes, categories, and themes, thereby enhancing the credibility and trustworthiness of the findings. The researchers will repeatedly review notes and audio recordings and follow these steps [30, 31]:

1. Carefully listen to the recorded interviews and transcribe them verbatim.
2. Read the transcripts repeatedly for a comprehensive understanding, identify meaningful segments, and perform open coding to create a codebook.
3. Apply the concept of the hermeneutic circle [30, 32] to interpret experiences related to device use, focusing on aspects such as convenience, regularity, motivation, and satisfaction.
4. Group similar meanings together.
5. Categorize meaningful sentences and commonalities, establish categories, and name them to derive sub-themes.
6. Re-examine the relationship between each interview’s context and the assigned categories for consistency.
7. Reassess the core meaning of the main themes and compile an overall care framework [32].
8. Integrate all content to produce a complete, holistic description

### 3.4 Data collection and management

All data will be pseudonymized. Unique IDs will be kept in a password-protected, encrypted network accessible only to the PI and study staff. Device fitting and troubleshooting will be documented, and adverse events will be immediately reported to the study team and logged in the participant manual. In our feasibility study, it is more crucial to ‘understand the reasons’ contributing to missing data rather than ‘deal with’ them. We will therefore describe the missing data and explore the mechanism in the semi-structured interview.

### 3.5 Safety monitoring

Participants may pause or stop device wear with any discomfort. Adverse events recorded with symptom descriptions and actions taken; referral pathways activated as needed. Serious adverse events (SAE) (e.g., hospitalization, ED visit, death) reported promptly and monitored; if three consecutive SAEs occur, the study will be temporarily halted.

### 3.6 Public and Patient Involvement (PPIE)

An Advisory Group composed of participating patients and caregivers will be developed to provide early feedback on device acceptability, environmental light exposure, and circadian measures to optimise feasibility for future relevant studies. Study results interpretation meetings and public education activities will disseminate findings in accessible language after the study is completed.

### 3.7 Study timeline

The project has enrolled its first participant on November 18^th^ 2025, and enrollment is expected to be completed in May 2026. Recruitment will continue until the last participating dyad has been confirmed, and data collection is plan to be done before July 2026. The final results are expected to be presented in December 2026.

## 4. Expected outcomes and discussion

Based on the research objectives, this study anticipates the following three merits:

1. Improving the application of wearable devices for patients with BPSD: By evaluating the acceptability and feasibility of Geneactiv and Re-Timer devices, this study will provide an abundance of information to help optimize their design and usage, thereby increasing the willingness of patients with dementia and their caregivers to use these devices regularly in daily life.
2. Collection of preliminary data on clinical efficacy: The study will generate initial data on the effects of Re-Timer on BPSD and sleep problems of AD patients and the stress and sleep on their caregivers. These findings will help assess whether the device can improve the quality of life for patients and caregivers and provide direction and baseline information for future large-scale studies.
3. Exploring the impact of light and circadian rhythm on BPSD: By analyzing long-term activity patterns and light exposure data collected via Geneactiv, this study will explore the relationship between light, circadian rhythm, and changes in BPSD symptoms. This may offer clinically meaningful insights for developing new non-pharmacological interventions and improving the assessment and management of BPSD.

Overall, despite its smaller sample size, this pilot study will be the first dyadic study exploring the feasibility of applying two wearable devices concomitantly on AD patients with pre-defined BPSD and their caregiver. Under the context of global ageing, the burden of dementia has not only led to huge caregiver burden, but also heightened societal and healthcare costs. If proved feasible in rural Taiwan where both digital and health literacy and resources are limited, our study may be able to guide future research in designing more customized wearable-based dementia care models that ameliorate BPSD and reduce caregiver burden.

### Ethical considerations

The study protocol adheres to the Declaration of Helsinki, and has been approved by the China Medical University and Hospital Research Ethics Committee with identifier: CMUH114-REC3-072. No compensation will be offered for the trial participants, however, if a participant experience harm that is considered directly due to the trial participation, the study team and the China Medical University Beigang Hospital will be responsible for the care and/or compensation.

### Trial registration

The trial has been pre-registered in ClinicalTrials.gov with identifying number: NCT07249918 (URL: https://clinicaltrials.gov/study/NCT07249918)

## Data Availability

n/a

## Data Availability

Upon study completion, we will share the minimal dataset required to replicate analyses (individual values behind reported means/SDs, raw device features, analysis scripts, metadata) upon reasonable requests.

## Author contributions

TWG and WJL conceived of the study and developed its hypotheses and analysis plan. LL and WFM supervised the development of the protocol. TWG, SHL, CSH, and CNC will collect the data, and TWG and WJL will analyse the data. TWG and WJL wrote the first version of the manuscript and sent it to all authors for review. TWG is the guarantor and attests that all listed authors meet authorship criteria and no others meeting the criteria have been omitted.

## Acknowledgments

We thank clinical staff, dementia trial managers (Yueh-Heng Chiang and Li-Yun Wan), and participating centers/hubs for recruitment support.

